# Does missing medication acutely change seizure risk? A prospective study

**DOI:** 10.1101/2025.06.06.25329144

**Authors:** Daniel M. Goldenholz, Joshua C. Cheng, Chi-Yuan Chang, Robert Moss, M. Brandon Westover

**Author notes:** Co-first authors (Daniel M. Goldenholz, Joshua C. Cheng). Corresponding author, 330 Brookline Ave, Baker 5, Boston MA 02215.

## Abstract

**Importance:** Medication adherence is widely emphasized in epilepsy management, with a belief that missing even single doses can trigger seizures. However, scientific evidence supporting this specific claim is limited, particularly regarding the impact of occasional missed doses rather than prolonged non-adherence.

**Objective:** To determine whether missing individual doses of anti-seizure medications (ASMs) increases short-term seizure risk in people with drug-resistant epilepsy.

**Design:** Prospective cohort study conducted over 10 months for each participant, with data collected from March 2022 through January 2023. Analysis was performed in February 2023.

**Setting:** Community-based study with participants recruited via emails and online services.

**Participants:** Adults with drug-resistant epilepsy experiencing 3 or more seizures per month, or adult caregivers of pediatric patients meeting these criteria. Participants used mobile applications to document both seizure occurrences and medication adherence.

**Exposure:** Daily documentation of ASM doses, with missed doses categorized as 0 (no missed doses), 1 (one missed administration period), or 2 (two missed administration periods) per day.

**Main Outcomes and Measures:** 27 participants (median age 29) were followed. The relationship between seizure occurrence and missed ASM doses was assessed using mixed-effects logistic regression models, controlling for baseline seizure risk using a 90-day moving average (the “Napkin method”). Various time windows for missed doses and analyses accounting for medication half-lives were examined.

**Results:** The Napkin method for seizure risk was significantly related to seizure occurrence (p<0.001). However, missed ASM doses the day prior showed no significant relationship with seizure occurrence (p = 0.68). This lack of association persisted when examining longer time windows of missed doses and when accounting for medication half-lives. Sensitivity analyses demonstrated that the statistical approach could detect even small effects (5% increased seizure probability) of missed medication, confirming that the negative finding was less likely due to methodology.

**Conclusions and Relevance:** While long-term medication non-adherence remains an important concern in epilepsy management, this study found no evidence that occasionally missing ASM doses significantly increases short-term seizure risk. These findings suggest that clinicians might reasonably reassure patients that infrequent missed doses may not dramatically impact immediate seizure likelihood, though consistent medication adherence should still be encouraged.

**KEY POINTS:** *Question:* Does missing individual doses of anti-seizure medications significantly increase short-term seizure risk in people with drug-resistant epilepsy?

*Findings:* In this 10-month prospective cohort study, the 90-day moving average of seizure frequency was significantly associated with seizure occurrence (p < 0.001), but missed anti-seizure medication doses showed no significant association with seizure occurrence (p = 0.68), even when accounting for medication half-lives or examining different time windows.

*Meaning:* While long-term medication adherence remains important in epilepsy management, occasionally missing doses may not significantly impact immediate seizure likelihood, which may help reduce unnecessary anxiety in patients who miss infrequent doses.

## INTRO

Protection against seizures is the number one concern for people with epilepsy. The mainstay of treatment is anti-seizure medication (ASM)^1^. The standard message to patients is that proper adherence to the ASM regimen is very important, because missing doses may precipitate seizures^2^, status epilepticus^3^ or death^4^.

But the scientific evidence supporting this standard message is scant. Specifically, there may be a difference between prolonged non-adherence and missing a dose or two occasionally. A recent review from our lab^5^ concluded that although short-term ASM nonadherence might increase seizure risk, there was insufficient evidence for a definite conclusion. Anecdotally, our clinical practice has numerous patients who admit to missing an occasional medicine with no subsequent seizure.

Our lab has demonstrated in several contexts that low-quality seizure forecasting can be achieved^6,7^ using a 90-day moving window average (the Napkin method) which calculates the number of seizure days over the past 90 days and divides by 90 to arrive at a predicted daily risk. The Napkin Method can be computed on a napkin. We found that it always performs at or better than chance permutation performance. This concept was validated in simulation, self-reported diaries and wearable device-based diaries^7^. For this reason, we see the Napkin method as a minimum bar for any clinically meaningful seizure forecasting^8^.

Therefore, we set out to obtain prospective data on a cohort of adults with drug resistant epilepsy, looking for evidence that missing a single dose of ASM can increase the risk of seizures. If missing a dose does have a significant impact on seizure risk, we hypothesize that forecasting with ASM adherence data will improve beyond the Napkin method.

## Methods

Following an IRB approved protocol (BIDMC 2022P000548), patients were recruited via emails and online services. Patients were screened for eligibility based on a diagnosis of drug-resistant epilepsy, a seizure rate of 3/month or more. Adult patients or adult caregivers of pediatric patients were encouraged to enroll. The study lasted for 10 months for each participant. Enrolled participants (or caregivers when appropriate) signed up with two free online services: Seizure Tracker and Medisafe. Both services provided an app on Android or iOS for mobile phone access. Seizure Tracker was used for 10 months of seizure diary recording. Medisafe was used for documenting every dose of ASM taken (or missed). Medisafe has optional medicine alarms as well as mobile alerts to document adherence for each dose. Once a week, the Medisafe app requested the answers to an online survey to confirm that the seizure diary is complete. Participants who did not complete the survey were contacted by study staff, and when possible completeness of data was confirmed. A de-identified data-table was generated based on the seizure dairies as well as the medication adherence data.

A mixed-effects logistic regression model was used to examine the relationship between seizure occurrence and missed ASM doses, and seizure risk as estimated by a moving average. A mixed-effects model was chosen as the data from seizure diaries were repeatedly sampled over time from the same participants, leading to non-independence of the data points. As such, the ability of the mixed effects model to account for intra-individual correlation within the data points was advantageous. Within the model, seizure occurrence was the response variable, whereas seizure risk as estimated by the moving average and missed ASM doses were the predictor (fixed effect) variables. The participant ID corresponding to each data point were used as the random effects variable.

To estimate the seizure risk for a given participant using a moving average, the number of seizure days was summed over a 90-day period and then divided by 90. This sliding window was then advanced by one-day increments and re-calculated, until the end of the window included the last time point. To generate the predictor variable for missed ASM doses, the number of missed administration periods (i.e., AM and/or PM dosing) was summated for each day. The output for each day and participant was a range from 0-2, where 0 = no missed ASMs, 1 = 1 missed administration period (either AM or PM), or 2 = 2 missed administration periods (both AM and PM). For the response variable of seizure occurrence, the output for each day was binarized as 1 = at least 1 seizure occurred that day, or 0 = no seizures that day. The mixed-effects logistic regression model was then used to examine whether the occurrence of seizures for a given day was related to the moving average of seizure risk from the prior 90 days, and/or missed ASM doses the day prior. Of note, as the very first moving average estimate required 90 days of lead-in data, the first datapoint for occurrence of seizures used was at day 91, and the first datapoint used for missed ASM doses was from day 90.

To determine whether the expansion of the 1-day time window would better reveal a relationship between missed ASM doses and seizure occurrence, the analysis was repeated for 1-day increases of the window up to 1 week prior. More specifically, for each iteration of window length, the number of missed ASM administration periods were summed for that prior time window and used as a regressor in the model.

To take ASM half-lives into account, a simplified analysis was conducted such that for each ASM, the maximum reported half-life for that medication was assumed. Furthermore, we assumed that if an ASM had reached at least one half-life due to consecutively missed doses, then that ASM was no longer “optimal” for that individual, even if still potentially therapeutic for them. To do this methodologically, when the number of consecutively missed doses of an ASM for a participant exceeded the maximum half-life for that ASM, then the subsequent time points were marked with a 1 (rather than a 0) as an indicator of increased seizure risk. This trail of 1’s would then continue until that medication was re-dosed. Of note, one additional advantage of using this approach is the lack of *a priori* selection of window length, as the effect of missed ASM doses utilizes the entire running dosing history up until that timepoint. The mixed-effects logistic regression analysis was then repeated with seizure occurrence as the response variable, with the prior 90-day moving average, and seizure risk the day prior as estimated by the half-life approach as the regressors.

Lastly, the sensitivity of the mixed-effects logistic regression analysis in detecting a possible relationship between seizures and missed ASM doses was examined. To do this, a simulated timeseries of seizures was generated for each participant based on their ASM dosing history. More specifically, in the day immediately following a missed ASM administration, the possibility of a seizure was generated with probability *p*. As such, if *p* was closer to 0, then the likelihood of a seizure the day after a missed ASM dose would be low, whereas if *p* was closer to 1, then seizure likelihood would be high. The mixed-effects analysis was then repeated at different values of probability *p*. The rationale here was that if the mixed-effects analysis was sensitive at detecting a relationship between missed ASM doses and seizures, then missed ASM doses would be a statistically significant predictor of seizure occurrence even when probability *p* was small (i.e., the effect of missed ASM doses on seizures was small). Given the probabilistic nature of this analysis when simulating seizure timeseries, the mixed effects analysis was also iterated 10 times at each value of *p*, with the mean regression coefficients and *p-value* calculated across the 10 iterations at each value of *p*. For the values of *p*, a broad range of values (0.01, 0.05, 0.1, 0.25, 0.5, and 0.75) were tested.

Our open-source analysis code is available at Github: https://github.com/GoldenholzLab/AdherenceStudy.git

## Results

A total of 27 participants were recruited and followed through the 10-month study. Basic information about seizure frequency, gender, and medications are available in the Appendix.

### The moving average but not missed ASM doses is statistically significantly related to seizure occurrence

Using the mixed-effects logistic regression model, the moving average as an estimate of seizure risk was statistically significantly related to seizure occurrence (*p* = 2.42 × 10^−57^). This relationship was positive (*β* = 5.47), such that higher seizure occurrence was related to a greater degree of seizure risk from the prior 90 days as estimated by the moving average. In contrast, missed ASM doses the day prior was not related to seizure occurrence (*p* = 0.68).

To assess whether the inclusion of longer window lengths of missed ASM doses can impact the relationship with seizure occurrence, the analysis was repeated with progressively longer time windows. However, whereas the moving average continued to be a statistically significant predictor of seizure occurrence, missed ASM doses were not a statistically significant predictor (Table 1).

**Table 1.** With incremental 1-day increases in window length up to 1 week, missed ASM doses were not a statistically significant predictor of seizure occurrence. In contrast, the 90-day moving average remained a statistically significant predictor of seizure occurrence in all these models (*p < 0*.*05*).

| Window length (days) | $\beta$ (missed ASMs) | $p$-value (missed ASMs) |
| --- | --- | --- |
| 2 | -0.13 | 0.43 |
| 3 | -0.07 | 0.53 |
| 4 | -0.01 | 0.94 |
| 5 | 0.03 | 0.69 |
| 6 | 0.03 | 0.60 |
| 7 | 0.02 | 0.69 |

### Even when accounting for ASM half-lives, ASM dosing is not a statistically significant predictor of seizure occurrence

The analysis was then modified to account for ASM half-lives. Using this approach, the moving average remained a statistically significant predictor of seizure occurrence (*β* = 5.49, *p* = 1.74 × 10^−61^). However, missed ASM doses were not related to seizure occurrence (*β* = 0.13, *p* = 0.63).

### The lack of relationship between missed ASMs and seizure occurrence is not due to poor sensitivity of the mixed-effects analysis approach

To determine whether the lack of relationship between missed ASMs and seizure occurrence was due to limitations in sensitivity of the mixed-effects approach, a simulation was performed whereby different seizure probabilities following missed ASM doses was generated. Even when simulated seizure probability following missed ASM doses was low (5%), the mixed effects analysis was able to detect a statistically significant relationship between missed ASM doses and seizure occurrence (Table 2). In fact, only an extremely low seizure probability following missed ASM doses (1%) led to a lack of relationship between missed ASMs and seizure occurrence using this approach (Table 2). Notably, the moving average remained a statistically significant predictor of seizure occurrence despite variations of seizure probability following missed ASM doses.

**Table 2.** Sensitivity analysis showing that the mixed effects regression model can detect a statistically significant relationship between missed ASM doses and seizure occurrence even when the probability of seizures following a missed dose is low.

| Probability of seizure | $\beta$ (missed ASMs) | $p$-value (missed ASMs) | $\beta$ (MA) | $p$-value (MA) |
| --- | --- | --- | --- | --- |
| 0.01 | 1.19 | 0.12 | 51.80 | $7.4 \times 10^{-3}$ |
| 0.05 | 1.99 | $1.16 \times 10^{-7}$ | 23.51 | $1.02 \times 10^{-10}$ |
| 0.10 | 2.24 | $4.99 \times 10^{-12}$ | 16.53 | $2.04 \times 10^{-13}$ |
| 0.25 | 2.77 | $6.74 \times 10^{-14}$ | 10.32 | $4.73 \times 10^{-25}$ |
| 0.50 | 3.41 | $7.85 \times 10^{-15}$ | 8.01 | $1.88 \times 10^{-44}$ |
| 0.75 | 3.80 | $2.51 \times 10^{-13}$ | 8.65 | $5.22 \times 10^{-23}$ |

## DISCUSSION

This study found evidence that there was no significant improvement over the benchmark Napkin method^7^ when adding information about missed doses of medications. To confirm that this finding was meaningful, we conducted a sensitivity analysis that found that even small impacts on seizure risk would have been identified due to the longitudinal nature of our dataset. We therefore suspect that more broadly, the short-term impact of missed medication is unlikely to have a dramatic impact on seizure risk.

This conclusion does not contradict other studies which explored the long-term impact of frequently missing medications^5^. In that case, it has been shown that poor medication adherence can increase seizures^2^, ED visits^4^, injuries^4^, status epilepticus^3^ and mortality^4^. Additionally, standard practice in epilepsy monitoring units (EMU) is to taper anti-seizure medications with the specific intention of changing seizure risk^9^. The fact that this standard practice is often successful further augments the clinical intuition that long-term medication adherence challenges increase seizure recurrence risk.

Conversely, it is well known among those who regularly work in EMU settings that the first missed ASM dose typically does not bring out seizures in most patients, supporting the findings here.

Hypothetically, missing a medication dose at “the wrong time” could have significant consequences. Specifically, it has been shown that there are daily and multi-day seizure risk cycles^10–12^. If a medication were missed during a period of higher seizure risk, perhaps this could represent “the perfect storm” and result in a seizure. It remains an effort for future studies to determine if this hypothesis is correct.

There are practical implications for this study. Many patients will miss doses of ASM occasionally, though this is often minimized if not asked in a permissive way^13^. It is recommended that patients be given psychological permission to admit missed doses^14^, and we have anecdotally found that asking “In an average month, how often do you miss a dose” results in very different answers (usually: “one or two”) compared with “Are you taking all your meds?” (usually: yes). It is generally good practice to gently remind patients about the safety afforded by proper ASM adherence, however the present study raises the possibility that “one or two” missed doses may be a reasonably acceptable answer to the adherence question.

There are several limitations to the present study. First, our primary data come from self-reported seizures and self-reported ASM adherence. Both were obtained in a curated manner-that is, involving weekly check-ins and oversight from the study team. However, it is well documented that self-reported outcomes such as these can suffer from over-reporting and under-reporting^15^, thus decreasing the certainty of the results. Our prior work^12,16–27^ suggests that there is often signal (biologically relevant data) despite the noise (distractors such as over- and under-reporting), and that self-reported data can be thought of as lower signal-to-noise ratio data compared with intracranial EEG.

Additionally, our study included a relatively small number of patients. The small sample size limits the generalizability of our results. We mitigated the small sample size somewhat using a long duration observation period of 10 months. Our models accounted for the different ASM half-lives being taken by our specific patients. Moreover, the seizure forecasting is well-known to be very challenging^8^. Therefore, it is certain that the choices of specific models described in the present study is included – we lack insight into what would be the “best” model for forecasting, therefore we defaulted to relatively simple linear mixed effects models that capture the essence of the Napkin method while adding simple covariates. We acknowledge that many other forecasting models are possible. By excluding simple linear combinations of these covariates, we aim to exclude “simple” possibilities. Future work with larger datasets may explore more advanced models.

In conclusion, despite the limitations above, we show evidence that short-term impact of missed ASMs does not appear to have a significant impact on short-term seizure risk. Despite this result, we emphasize that there are numerous reports that long-term impacts of medication non-adherence are dangerous and would always recommend medication adherence whenever possible.

## Data Availability

All data produced in the present study are available upon reasonable request to the authors

