## Appendix for "Does missing medication acutely change seizure risk? A prospective study"

Basic statistics of study population:

Age Statistics:
Minimum Age: 6.0
Maximum Age: 77.0
Mean Age: 33.2
Median Age: 29.0

Number of patients: 27
Percentage of Female Patients: 33.3%

Seizure Rate Statistics (per month):
Minimum Seizure Rate: 0.11
Maximum Seizure Rate: 76.26
Mean Seizure Rate: 11.26
Median Seizure Rate: 4.06

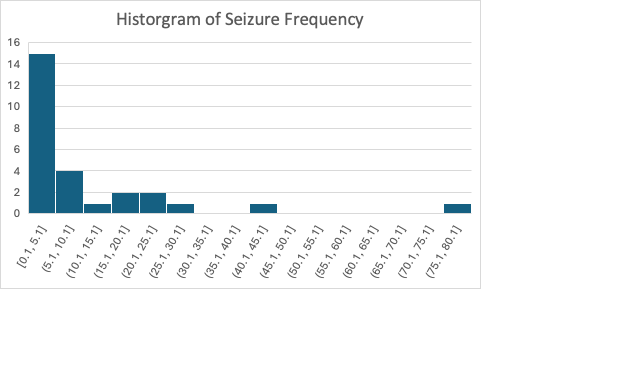

Medications taken by study patients:

brivaracetam

carbamazepine

CBD

cenobamate

clobazam

clobazam

eslicarbazepine

felbamate

fenfluamine

lacosamide

lamotrigine

levetiracetam

oxcarbazepine

perampanel

phenobarbital

rufinamide

topirimate

valproic acid

zonisamide
